# The policy formulation process, and the role of actors in the policy formulation and implementation process: A policy analysis of the Kenyan free maternity policy

**DOI:** 10.1101/2024.01.26.23300268

**Authors:** Boniface Oyugi, Zilper Audi-Poquillon, Sally Kendall, Stephen Peckham, Edwine Barasa

## Abstract

Kenya is one of the many African countries committed to advancing its health system reforms by providing affordable and equitable access to essential health services. In 2016, the Government of Kenya unveiled an expanded free maternity care policy called *‘Linda Mama’* to provide essential health services for pregnant women. We explored the agenda setting and the formulation of this policy to understand the processes, content and context, and the role of the actors in the formulation and implementation. We conducted an exploratory qualitative study, which involved document review, key informant interviews (KIIs) with national stakeholders, and in-depth interviews with County officials and health care workers (HCWS). We used a theoretical framework capturing the preliminary situation analysis of the policy, the processes, the content, and the stakeholders’ roles in the formulation and implementation. This study was conducted in three facilities (levels 3, 4, and 5) in Kiambu County in Kenya. Data were audio-recorded, transcribed and analysed using a framework thematic approach. We found that the priorities of the policy were mainly to meet a political campaign agenda but also align them with the objectives of the country’s legal and policy-guiding instruments and the global goals of SDGs that sought to improve the quality of maternal and neonatal care and eliminate financial barriers. The policy also sought to enhance access to skilled birth attendance (SBA), and its redesign filled the challenge of the previous policy. The stakeholders (bureaucrats, professional bodies, public and developmental partners) influenced the processes of the formulation and the content of the policy through their power to put forward and advocate for specific ideas through issues framed in a political and socioeconomic context. Several stakeholders played different roles in the formulation and implementation based on their interests, power and position in the ecosystem of the policy. Policy formulation or change requires the agents to work within the relevant context, stakeholder interests, power, ideas and framing of issues.

## Introduction

Kenya is one of the many African countries committed to advancing its health system reforms by providing affordable and equitable access to essential health services [1]. For instance, user fees were introduced in public health facilities in the 1980s, and those for outpatient care were suspended in 1990 because of equity concerns but later reintroduced in 1991 [2]. Free maternal deliveries were introduced in all public healthcare facilities in 2007, but its formulation and implementation was not well documented [2]. In 2013, following a presidential policy directive, Free Maternity Services (FMS) was introduced in all public health facilities [3, 4]. The National Government financed the FMS policy with the funds paid directly to healthcare facilities at Ksh2500 (approximately 27 USDs as of the implementation time) for every delivery in primary healthcare facilities, whereas the sub-county hospitals were reimbursed KSh5000 (approximately 55 USDs) for every delivery, normal or caesarean [1]. However, it faced implementation challenges, specifically with regards to poor service delivery, due inadequate preparation before its rollout [5]. Thus, in October 2016, the National Government unveiled an expanded free maternity policy called ’Linda Mama’ [Swahili word for - take care of the mother], managed by the National Hospital Insurance Fund (NHIF) to address the challenges from the previous policy [6, 7]. It provided a package of essential health services for pregnant women accessed by all in the targeted population based on need and not the ability to pay [7]. Linda Mama aimed to achieve universal access to maternal and child health services and contribute to the country’s progress towards Universal Health Coverage (UHC).

Since its implementation, several studies have reviewed the Linda Mama programme, but have only focused on the implementation, its effects and impacts, quality of care and the cost [2, 6, 8–13]. To our knowledge, no study so far, has documented this policy formulation process, and there remains a dearth of knowledge in this area. Yet, researchers have shown how problems with policy execution could be enhanced by having a joint consideration of policy design (sometimes defined as the crafting of comprehensive causal assumptions, goals and visions, rules, tools, strategies and organisations to address a particular policy problem) and policy execution. Policymakers create policies that deliver desired results by carefully aligning the problems, solutions, interests and organisational resources by properly working on the policy design, which postulates plausible scenarios and anticipates future implementation problems [14]. Further, policy formulation and agenda setting are essential fields of enquiry which can give insights into the complex formulation process to show how getting and maintaining policy issues on the agenda is a crucial part of decisions made during policy development and implementation [15]. Most policy development or formulation (whether as an intent, a written document, or a practice) does not often follow a particular format as it is a complex and intertwined process [15, 16]. It is difficult to predict.

Researchers have shown why some issues make it onto the policy agenda while others fail. For example, some have argued that the structure of organisations could explain why some issues are considered and have an advantage vis-a-vis the alternatives [17]. At the same time, individuals’ or even institutional processes could influence what is to be addressed at any given time. Others have emphasised the role of external events or public opinions, and how these two combine with political incentives to shift attention to the policy agenda [17]. Some issues are likely to get a better response from stakeholders if they have high legitimacy, are highly feasible, and have high support [15, 18]. Besides, the framing of the problem is equally important as it influences how policymakers tackle it. Noteworthy is the strength of the organisation and individuals concerned with the issue, and how those involved understand and portray it [19]. Finally, the features of the problem (issue characteristics) and the political context (the environment in which the actors operate) also play a role [19].

Kenya has made some good progress towards reducing neonatal and mortality rates and improving the coverage of health services in the past decades. For example, the maternal mortality ratio (MMR) decreased by approximately 6% (from 564 in 2000 to 530 in 2020),[20] while the neonatal mortality rate reduced from 33 to 21 deaths per 1,000 live births between 1990 and 2022 [21]. However, despite this progress, many women, neonates and children in the country still experience morbidity and mortality from preventable pregnancy and child health-related causes [22]. There has also been an increase in the live births assisted by a skilled provider over the past two decades, from 41% in 2003 to 89% in 2022 [21]. This increase has been attributed to the free maternity policy that the government introduced in June 2013 [23].

This study, therefore, seeks to advance the understanding of Kenya’s free maternity policy (FMP) agenda setting and the policy formulation process. It provides some evidence of the issues that got into the policy formulation agenda (agenda setting and selection of alternatives), and how they got there. It also explores the role of the actors in the free maternity policy process.

## Methods

### The guiding conceptual and analytical framework

This policy analysis is based on a conceptual framework derived from literature review and health policy analysis frameworks (Table 1) [24]. It captures the background of the policy, derived from Hercot et al.’s [25] work, which focuses on the preliminary situation analysis and setting the priorities of the policy (an understanding of the origin of the policy). It also emphasises the portrayal of an existing window of opportunity needed to restrict the ‘inventory phase’ of a policy [26]. It then draws the policy formulation process from Walt and Gilson’s [27] policy triangle of *actors* (whose roles, power, and influence during formulation and implementation was analysed through a stakeholder’s analysis [28–30]) following into its application in the SHIELD project,[31] *processes* involved during formulation, *context* (political, social and economic, local and nationally) of the policy, and *content* (envisaged design) of the policy formulation [15].

**Table 1:**
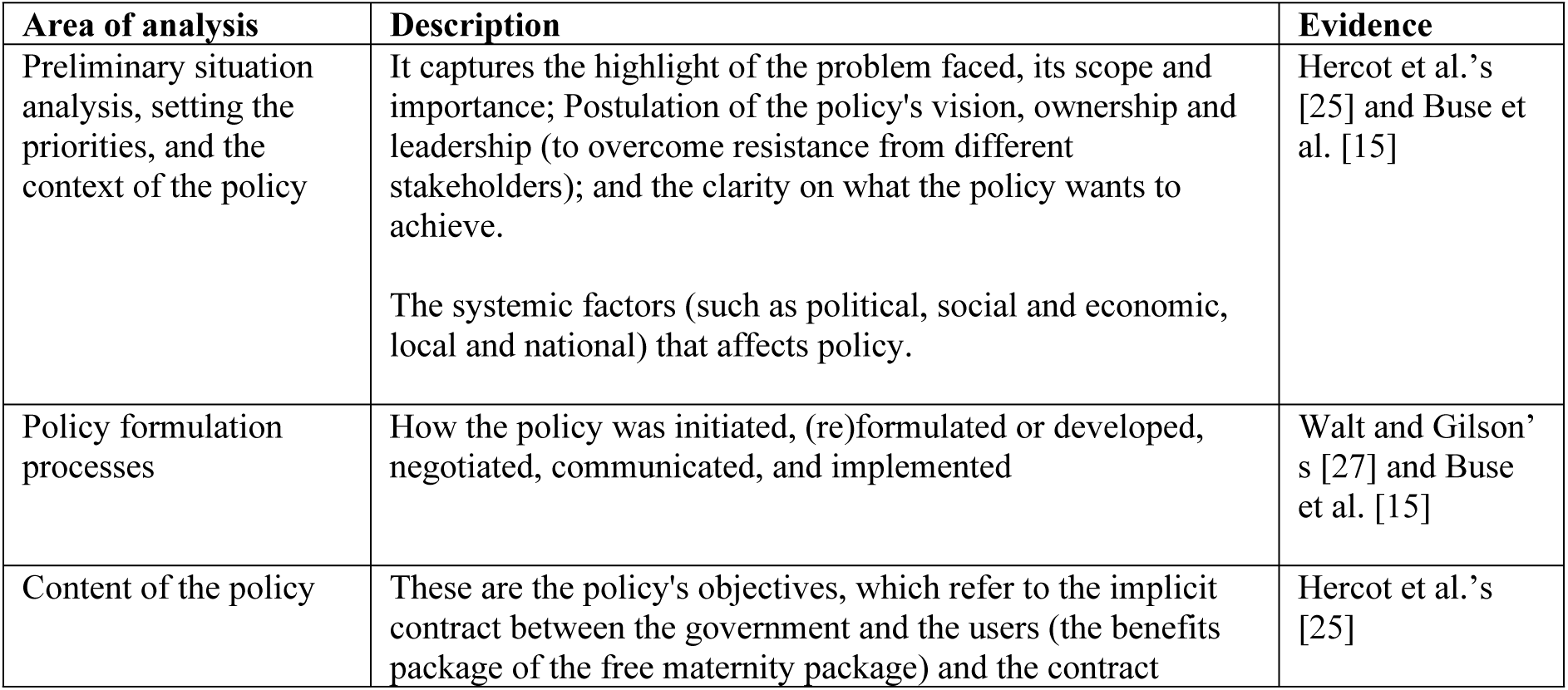
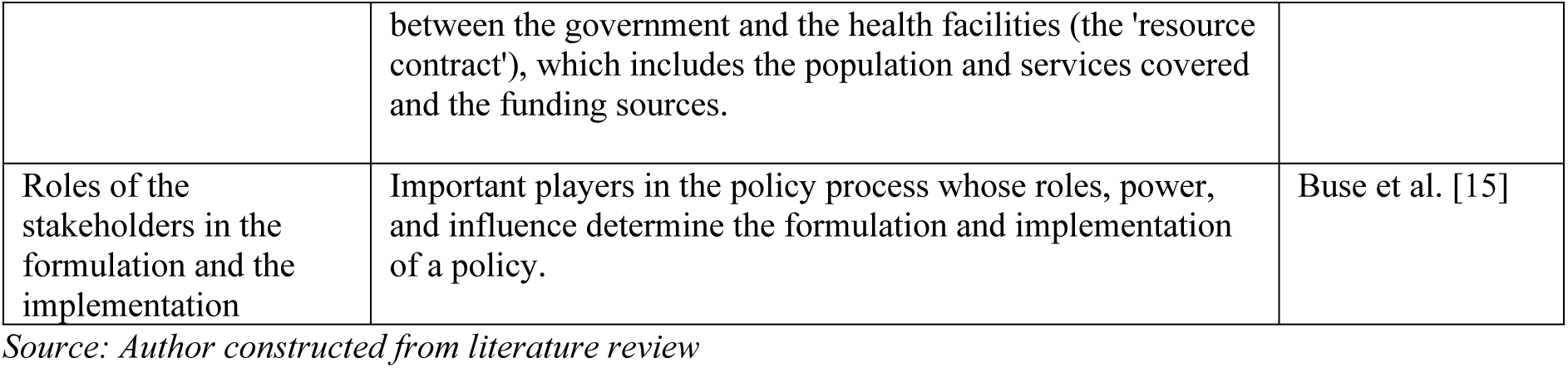
Guiding framework for evaluating the policy process.

| Area of analysis | Description | Evidence |
| --- | --- | --- |
| Preliminary situation analysis, setting the priorities, and the context of the policy | It captures the highlight of the problem faced, its scope and importance; Postulation of the policy's vision, ownership and leadership (to overcome resistance from different stakeholders); and the clarity on what the policy wants to achieve.<br>The systemic factors (such as political, social and economic, local and national) that affects policy. | Hercot et al.'s [25] and Buse et al. [15] |
| Policy formulation processes | How the policy was initiated, (re)formulated or developed, negotiated, communicated, and implemented | Walt and Gilson's [27] and Buse et al. [15] |
| Content of the policy | These are the policy's objectives, which refer to the implicit contract between the government and the users (the benefits package of the free maternity package) and the contract | Hercot et al.'s [25] |
|  | between the government and the health facilities (the 'resource contract'), which includes the population and services covered and the funding sources. |  |
| Roles of the stakeholders in the formulation and the implementation | Important players in the policy process whose roles, power, and influence determine the formulation and implementation of a policy. | Buse et al. [15] |

*Source: Author constructed from literature review*

### Study Design

This study utilized an exploratory qualitative design which involved document review, key informant interviews (KIIs) with national stakeholders, and in-depth interviews with County officials and health care workers (HCWS).

### Study setting

This study was part of a larger study that sought to examine the policy process, quality and cost of free maternal healthcare in Kenya [7]. We therefore purposively selected Kiambu County for our study, which was conducted between November 2018 and June 2019. Kiambu County was chosen because of the logistic feasibility of data collection and the sociodemographic characteristics, health indicators and population size [32–34]. It is the second-most populous county in Kenya after Nairobi City County, with a population of 2,417,735: 49.1% male and 50.59% female [32] 26.9% of the population in Kiambu are female of reproductive age (15-49 Years),[33] and 89.2% of births in the county happen in a health facility and 98.2% of births provided by a skilled provider [21].

### Study population and sampling

We conducted key informant interviews (KIIs) with national level policymakers (n=15); and in-depth interviews with sub-national level/County policymakers (n=3); and facility level decision-makers (n=18). All these informants were purposively selected based on their knowledge of the FMP, and included officials drawn from the Ministry of Health (MoH), the national health insurance agency (NHIF); development partners; County department of health officials; facility-in-charge and HCWs and others. Documents reviewed included legal documents (n=7), websites (n=5), and other documents (n=8) as described in Table 2.

**Table 2:** Summary of respondents and documents reviewed.

|  | Female | Male | Total |
| --- | --- | --- | --- |
| **County-level and health facility respondents** |  |  |  |
| The county department of health officials (senior-level and middle-level managers) | 2 | 1 | 3 |
| Public hospital managers (Medical superintendent, facility-level managers, department in-charges, hospital administrator) | 7 | 2 | 9 |
| Health Workers (nursing officers, clinical officers, accounting/ clerical officers) | 6 | 3 | 9 |
| **Total** | **15** | **6** | **21** |
| **National level respondents** |  |  |  |
| Ministry of Health officials | 4 | 1 | 5 |
| NHIF officials | 3 | 0 | 3 |
| Development partners | 2 | 5 | 7 |
| **Total** | **9** | **6** | **15** |
| **Documents Reviewed** |  |  |  |
| **Category** | **Item** |  |  |
| Legal documents | 1. The Constitution of Kenya (Article 187, Article 43)<br>2. The Health Act (Article 2, Article 5, Article 6, Article 7)<br>3. National Hospital Insurance Fund Act<br>4. The County Governments Act, 2012<br>5. Intergovernmental Relations Act, 2012 (section 25)<br>6. Legal Notice 137–183 of August 2013 (which exercise the powers conferred by section 23(1) of the Transition to Devolved Government Act, 2012)<br>7. Legal Notice No.34 National Government Regulation |  |  |
| Websites | 1. The Standard Group PLC<br>2. The Nation Media Group<br>3. Kenya Ministry of Health<br>4. The World Health Organisation<br>5. The World Bank |  |  |
| Other documents | 1. Linda Mama implementation manual<br>2. NHIF PowerPoint presentations on Linda Mama (2)<br>3. Draft free maternity service policy<br>4. Rapid assessment of Linda Mama report<br>5. Other Linda Mama case studies reports (2)<br>6. Facilities maternal and child health progress charts and wall hangings/ posters |  |  |

### Data collection and analysis

We conducted in-depth key informant interviews (KIIs) with the national level respondents, and in-depth interviews with the implementers (from the county and the facilities), using semi-structured interview guides developed to capture the formulation processes and implementation experience of the FMP. We first piloted the interview guides in a non-participating facility, to ensure the questions captured all the aspects of our research objective. The interviews were conducted in English and audio-recorded, each lasting between 30-60 minutes.

All the interviews were transcribed verbatim in English and compared against their respective audio files by BO for transcription and translation accuracy. All the validated transcripts were imported into NVivo 12. The data were analysed thematically [35–37] following the conceptual framework described prior. One researcher (BO) assigned unique identifiers to the data, familiarised himself with the data through immersion and repeatedly read and re-read the transcripts. He then started by developing ‘lower-order premises evident in the text’ [38] through open coding (assigning codes to portions of data) [39], thereby creating an initial coding framework. Study team members (SK and SP) reviewed and discussed the initial coding framework, and any discrepancies were appropriately reconciled. The final coding framework was applied (by BO) to the data and later charted the data to allow themes to emerge through comparisons and interpretations.

### Ethical consideration

Ethical approval for this study was obtained from the University of Kent, SSPSSR Students Ethics Committee and AMREF Scientific and Ethics Review Unit in Kenya (Ref: AMREF – ESRC P537/2018). Further, we received permission to conduct the study from all the healthcare facilities where the study was conducted and additional clearance to conduct research from the County Government of Kiambu, Department of Health Services (Ref. No: KIAMBU/HRDU/AUTHO/2018/10/31/Oyugi B). We also got written informed consent from the respondents before conducting the interviews, after informing them about the purpose of the study and their right to withdraw consent at any point. They were also assured of their confidentiality, and that their data would be reported in an aggregated format, and anonymised to protect their identities, throughout the course of this study.

## Results

### Preliminary situation analysis, setting the priorities, and the context of the policy

The preliminary situation analysis has revealed that the priorities of the policy are linked to several factors, such as the desired needs of the government and key stakeholders and the need to bridge the gaps from the previous FM policy. These aspects set the context of the policy’s adoption and implementation.

#### The government prioritised the FMS policy to reflect and align with the objectives of the country’s legal and policy-guiding instruments

The government sought to align and improve the coverage of reproductive maternal and neonatal child health (RMNCH) services as reflected in the Constitution of Kenya, 2010, the Vision 2030 and the Health Sector Strategic and Investment Plan (HSSP) 2019-2023 [40]. For example, as noted by the respondents, the constitution provided every citizen with the right to quality health and life (including maternal and neonatal care) [41] while the social pillar of the Kenya Vision 2030 envisioned a country-wide scale-up of community health high-impact interventions to strengthen and enhance the implementation of level one MNCH high-impact intervention services (such as free maternity service) to accelerate initiatives targeting nutrition services, family planning, immunisation, sanitation and safe motherhood [42]. Besides, the HSSP envisioned increasing equitable access to care regarding quality and availability of services at all levels and creating and sustaining demand for improved preventive and promotive healthcare services [43].

#### The need to align with the global goals of achieving sustainable development goals (SDGs)

Achieving SDGs was seen as a key tenet and foundation that anchored the need for the FMP agenda. It was noted that while Kenya had made some progress in achieving the Millennium Development Goals (MDGs), the country had missed those targets [44]. Therefore, the policy was triggered by the gaps in those goals, as well as the desire to achieve the maternal and child health targets in the SDGs:

> *‘…As previously we had been given the MDGs [Millennium Development Goals], but they didn’t work for the 15 years. So now we are working on the SDGs [Sustainable Development Goals]. So being a healthcare worker we are trying our best to ensure that the country and our county achieves its objective’ – **(R003, Nursing Officer)***.

#### The need to improve the quality of maternal and neonatal care, by eliminating financial barriers, thereby enhancing access to skilled birth attendance (SBA)

There was an acknowledgement that the enhanced FMP was an incentive to boost the quality of maternal care, since most women had inadequate access to SBAs. As noted by the respondents, the Kenya Service Availability and Readiness Assessment survey had showed the existence of sub-standard care in facilities, and therefore, the enhanced policy was meant to boost maternal and neonatal health, that had deteriorated in quality within the facilities. In addition, there was the need to eliminate financial barriers which would enhance access to free maternity services by pregnant women. As perceived, the FMS policy would address concerns of other barriers of access; such as geographical barriers (ensuring that the marginalised populations and those in rural areas have equal access); socio-cultural barriers (ensuring that the use of traditional birth attendants (TBAs) reduces; and service access (ensuring access to services such as testing at ANCs which were previously unavailable), thereby enhancing the achievement of UHC:

> *‘Linda Mama also informs Universal Health Coverage, because Universal Health Coverage seeks to enhance, seeks to embrace access to health care providers.’ – **(R034, NHIF Officer)***.

#### The desire and urgency to achieve a political campaign agenda

Some respondents perceived the FMS policy to be a political tool used by the government to fulfil a campaign agenda, as captured in the president’s *Jubilee Party 2013 Presidential campaign Manifesto* [45]. Some of the respondents highlighted that the goal of the policy was to fulfil part of the Big Four agenda that the then president formulated after his election, and policy makers and implementers had no option but to implement it:

> *‘it was used as a political tool…to be able to achieve and acquire power…I can tell you in terms of even conceptualizing the idea implementation, politics played its part, and I think the president announced it in 2013 during, is it Madaraka day [Kenyan public holiday]…so there is always politics behind some of these things…I think it was politically appearing so usually the health workers have no…choice but to actually actualize’ – **(R026, Development partner)***

#### The need to include the private sector as the additional implementers of the policy

There was the need to rope in the private sector and faith-based organisations (FBOs), who were not implementers in the previous FM policy. The move was aimed at decongesting public facilities and giving pregnant women more choice. Further, the move was thought of as being imperative in improving access in areas, especially arid and semi-arid areas, where there were not many government facilities, but that had many FBOs offering services. Eventually this would end up improving efficiency:

> *‘….we weren’t including private, and as you know private almost takes care of 40% of our population, only 60% more often that uses GOK [Government of Kenya]…there was a feeling that we are leaving a few people behind, especially in Nairobi, where we have more private facilities than public….so that was one of the driving forces, FBOs [Faith Based Organisation] also thought left behind, who are our partners in so many ways….There was also the consideration of the far front [marginalised] areas where there are private and mostly even FBOs and there is no GOK facility around, but there is a lot of FBOs especially in Turkana, so it was thought [important] for us to improve on access and efficiency’ – **(R032, MoH Official)***.

#### The need to have uniformity in the delivery of the maternal service provision under the free maternity policy

It was perceived that the policy, as implemented, needed to have uniformity of service provision across the counties (especially those that had launched their own unique service delivery platforms for their citizens). This would allow citizens from a different county to access services in another county even if they hadn’t registered there. It would enhance competition amongst counties and reduce the incentive to seek healthcare services in counties that had invested more in their healthcare services:

So, I guess the structure levelled out the advantage that already developed counties would have in terms of service delivery to their citizens. Because all we need is a card or a registration, it doesn’t matter which county you came from, and you will be able to access your care from whichever county you go. That is very different from the roll-out you see in other counties, where a county is rolling out its own program and you can only get that care if you are from [reside? in] that county and are registered as a member of that county. That discrimination also ruled out the advantage of having citizens access care in neighbouring [counties] and not pay for it [as] somebody else pays for it.’ – **(R010, Facility manager).**

#### The policy was to cover the loss of funding that was previously charged to the mothers for the services, but that would no longer be available

The facilities needed to continue sustaining the health facilities costs. However, it was also linked to the need to improve the quality of care from the reimbursements to the facilities (incentivise the facilities). It was noted that the funds would help purchase equipment that the government had been unable to provide, motivate staff through incentives, and employ additional staff:

> *‘You know cost sharing; in the past you know, maternity used to generate cash and now it’s zero because it’s free and services have to run…I think it’s purely because of financial reasons’ **– (R004, Nursing Officer)***.

#### The policy was addressing the challenges of the previous free maternity policy

The implementation challenges experienced in the previous FM policy were perceived as the substantial reason for the shift to the current *Linda Mama* policy. The respondents at both the national and the county level referred to the challenges that had been noted by Tama et al. [5] in their process evaluation process evaluation as: *lack of exhaustive service package* due to inadequate costing of the services; *data problems*, where facilities were using inflated utilisation numbers, rather than mothers’ unique identifiers, to get claims from the MoH that were unverifiable; *poor quality of care,* since patients was pushed to the public hospitals that were ill-prepared to tackle the high number of mothers because of inadequate infrastructure. Additionally, there was a *lack of or inadequate communication* of the policy at the grassroot level, which led to poor clarity of the content of the policy, and *disappointed and dissatisfied clients* with the services. The respondents noted that the work was overwhelming to the MoH Department of policy and planning, and reproductive health, which did not have the capacity to manage both the payments and the services:

> *‘Data fraud, issues about data verification, data validation to be able to monitor the utilization rate, second the issue of disbursement of money, proper disbursement of money from the Ministry of Health to the health facilities, thirdly is the fact that private sector was left out.’ – **(R029, Development Partner)***

### The processes of formulation of the free maternity policy

#### The private sector network had a significant interest in the policy formulation phase

The justification of the inclusion of the private sector by the private sector networks and negotiation on the pricing characterised the discussions at the formulation stage. For instance, in using previously conducted research, the networks observed that the developed systems in the private sector would ease the previous FMP implementation challenges in the public sector. As such, the networks opted to push the private sector agenda of their inclusion through the national and county leaderships (using the success of the programs they had implemented prior, such as leveraging their network of community health volunteers which would make them favourable when the policy was finally rolled out). Besides, through their established interests and strengths, as soon as the policy had been formulated, they worked with their networks of private health facilities for advanced accreditation using internal quality of health standards and guidelines before the actual Linda Mama accreditation. They also communicated the packages of the policy in advance to their network of hospitals:

> *‘…we gave our input at the National level, but also, we realized that we needed to do some groundwork at the County level, because these are two independent Governments…so at the MOH level, so we went out to the CHMT’s [County Health Management Team] at the community level and just made them understand what we are trying to do and why this is important and why they should support us, for them to be able to deliver on free maternity care through the private sector. And so, what we saw the CHMT’s do is that they attached themselves to our teams and they did routine supportive supervision with us on a sampling basis just to ensure that what we were telling them was what was on the ground’ – **(R030, Development partner)***.

The perspective from the private sector network was that the NHIF could not manage (lead) the extensive quality of service that the private sector, the Non-Governmental Organisations (NGO) and the Faith Based Organisations (FBOs) had achieved so far because their reimbursements were perceived as unattractive. The NGO and FBOs were keen to join the policy scheme but proposed that the government raise the reimbursement cost. The concern was that their investment in infrastructure and other additional costs, such as rent and staff, was high:

> *‘Within our…400 plus providers….NHIF cannot manage that quality…you see there are so many things, it’s [the policy] creating access, and it’s creating equity in term of it reaching the poor…not so many people appreciate it because it’s not attractive in terms of profit but there are a lot of private sectors that are actually improving on that… we are not saying you put it at 10000/= [USD 95.9], we are saying put it at 6000/= [USD 57.5] there will be demand and they will increase. And then contract other facilities to support you, contract other quality organizations like PSI, MSI who are known for quality in terms of SRH to support the Government in terms of quality.’ – **(R029, Development partner)***.

#### The costing of the policy at the formulation stage was done using the projections of maternal indicators and funding ascertained

The policy was conceived as an insurance package allowing mothers to use the registration card, guarantee maternal care for up to a year after birth and transition to an NHIF card for outpatient and inpatient services at their facilities of choice. The three bases of costing at the formulation were that a) health is a free good, b) the need for introducing sustainability in developing the provision of maternal care, and c) the expected number of deliveries given the demographic characteristics of the mothers. It was estimated that there would be almost 1 to 1.2 million mothers delivering in the country every year, with a conservative estimate of 25% being insured by entities other than NHIF. Eventually, the FM policy would reach 700,000 mothers by the first year of roll-up, who, after having utilised the services and seen the value of it, would be attracted to it as some form of insurance. Consequently, it was further envisaged that 25% of those who use the free policy services, especially those in quantiles 1 and 2, could transition to full insurance after the expiry of *Linda Mama* and start paying for it. The formulation idea was that – in the following years of the policy – with the mothers in quantiles 1,2, and 3 paying a monthly contribution fee to the NHIF for themselves, it would ease pressure on the treasury fund meant for the policy. Additionally, there was a projection of the prospective number of both vaginal and caesarean deliveries that was envisaged, with the assumption that 15-20% of the deliveries would complicate:

> *‘So, all those were projected, the caesarean section how many do were expected, close to 10%, 15%, the normal ones…’ – **(R032, MoH Official)*** and *‘…when the fund was being put in place, it was assumed that at least fifteen to twenty percent would complicate [result to a complication]. So, they will be catered for by everything else.’ – (**R024, MoH Official)***.

The overall projection cost of the policy was estimated at KES 6.5 billion (USD 62.3 million) (Exchange rate, 1 USD = KES 104.32, which was the rate as of 1^st^ January 2017 at the initiation of the first phase of the implementation of policy (from https://www1.oanda.com/currency/converter/), but the national treasury allocated the MoH KES 4.2 billion (USD 43.1 million) which the respondent deemed sufficient to meet the need.

Further, the funding source for the policy was ascertained. Unlike the previous policy, co-funded by the development partners (JICA and The World Bank), the *Linda Mama* policy was mainly conceived as tax-funded by the national government. However, the respondents noted that there was no specifically earmarked funding for the policy.

> *‘It is largely funded by our taxes…and they are not earmarked, so that means that they could easily be rerouted somewhere else.’ – **(R023, MoH Official)***

#### There was general perception that the costing had not been done in a consultative manner as should be the norm

While the majority of the national-level stakeholders perceived [argued/felt] that the policy had been sufficiently costed including assessing its sustainability, others felt that the program had been arbitrarily costed and had made many costly assumptions:

> *‘…we cost it according to what we spend and then we put an arbitrary amount for miscellaneous, we are not thinking of what is the cost-benefit of this like can you relate every cost you put in, like for a hundred bob we put into healthcare, how many maternal deaths are averted…we don’t know why; we cannot justify why we put a mark of four billion at least scientifically’ – **(R023, MoH Official)***

Consequently, the respondents at the meso level felt that the costing was somewhat dubious based on the implementation experience that had left them with more costs to absorb from the service provision:

> *‘You don’t know how the figures of reimbursement were arrived at and noting that every county is a market in its own way, with its own influencers of supply, demand and price. Giving one cut line of a price disadvantages those counties where services are provided at a higher cost compared to counties where services are provided at a lower cost. So, for you to remain in the system you had to absorb some cost and absorbing the cost [would mean] you have a loss. And again, with coming to be like a revenue loss for the county to keep on reimbursing services for which their people are asking for but for which the money funding is not adequate per case.’ – **(R010, Facility incharge)***

#### NHIF was chosen as the ultimate purchaser of services

at the formulation stage, the NHIF was the agency proposed to run the FM fund, as it had the technical and institutional capacity to manage such funds based on its experience. There was no other consideration of transitioning the policy to other organisations other than the NHIF. The majority of the respondents believed that NHIF was undergoing reforms at the time aimed at transitioning to UHC; hence, had better accountability structures. Additionally, NHIF had existing structures and networks of facilities across the country that could be leveraged instead of creating a different program. Besides, NHIF had already been working with the private sector, hence, it was easy to attract and enroll many facilities. The plan, therefore, was to utilise NHIF instruments to track elements such as average length of stay, quality of care, access, fraud, complaint system and payment of providers:

> *‘No there were no other options…[be]cause [with] Linda mama…we wanted really to involve the low-cost privates to improve the access to mothers which we would not have done as MOH [be]cause MOH can’t pay money to private but the law allows NHIF to pay to do that. So that was one of the driving force[s]. NHIF can pay private I mean they have contracts with them, so it was easy we weren’t going to start something new’ – **(R032, MoH Official)***.

While some respondents felt that there were no legal hurdles in working with the NHIF – a semi-autonomous agency under the MoH – document review revealed that civil society organisations had been pushing the narrative of the illegality of the process (see, KELIN Kenya [46]). Additionally, engaging the NHIF would remove the challenge of returning unconsumed money to the treasury as was the norm when the MoH managed the program during the previous policy. The process would allow for exhaustive use of the finance allocated for the project in the rolling years:

> *‘So, what happens when the money is within the ministry of health, when the financial year comes to the end, that money goes back to national treasury, through the process, the government budgeting process and it is availed again the following financial year. So in such a scenario where now it goes to NHIF such a body corporate, that money does not have to go back and perhaps if there are pending reimbursements like for instance if I am doing reimbursements for the last quarter of the financial year, April, May, June, and you see like for end of June or end of May, the facilities have to report, then it is compiled, then it is paid, which means it will be paid post, the financial year is ended and the money will already have gotten back. So, they won’t be paying for that period. So that kind of challenges they required an institution that can be able to handle that’ – **(R024, MoH Official)***

#### The implementation manuals and guidelines were agreed upon at the formulation stage but there were gaps in the monitoring the resulting quality of maternal care from the policy

The formulation committee developed a policy document and a concept note which were eventually taken to the Cabinet for approval. In addition, an implementation manual and a communication strategy were also developed.

To kickstart the process of implementation, a memorandum of understanding (MoU) between the MoH and the NHIF was signed. Correspondingly, financial guidelines were also developed but it was envisaged that the HCWs would rely on the clinical and service provision guidelines that were in use prior to the policy. Notwithstanding that the MoH, through the department of quality standards developed the Kenya Quality Model of Health (KQMH) to provide structured support to counties for general QoC, there was no other guideline on quality maternal care developed at the formulation of the policy, as noted by respondents. There was a feeling that the little attention was being paid to quality guidelines but more on the implementation. The private organisations and development partners proposed to fill up the gap in quality through their own project such as *safe care* which trains and teaches the staff in the facilities on improvement of quality:

> *‘MOU with the ministry whereby we looked at how to bring in a bit of technology to assist in the registration of women so that through NHIF system we are able to register first, once you register, the next step is that there has to be a confirmation of pregnancy. So, we signed an MOU on what will be our roles in line with the implementation of this product’ – **(R025, NHIF Official)***

> *‘…not squarely on the government, but thanks to development partners and private entities…there are very clear projects or approaches that come into address some of these gaps that are identified. So, I say the facilities are willing, but it only works where they have development partners’ – **(R035, Development partner)***

### The Content of the policy

#### The envisaged design envisioned inclusive benefit packages, more infrastructure, and human resources

The envisioned package cuts across maternal care from ANC, delivery, to PNC care, complications and referral services. Furthermore, it was envisaged to take care of the infant within the one-year period in the program (see Fig 1). Further, at the formulation, it was projected that the workload would increase, requiring more investment in infrastructure and human resource:

> *‘We did not go into those details but we also said as a, the government need to invest on human resources, we are anticipating some increases in human resources, I mean increases in workload, I think that should be followed by investment in human resources, investments in commodities and also even infrastructure, other infrastructures, renovation of the maternity wards, those were things that we had anticipated and we recommended investments in those areas as to whether that happened is a different issue.’ – **(R026, Development partner)***

As further noted by ***R026, Development partner***, *‘…the transport element was not factored in’* at the formulation hence mothers would spend money on transport. At the formulation stage, the envisioned reimbursement of the scheme was as shown in Fig 2.

**Fig 1:**
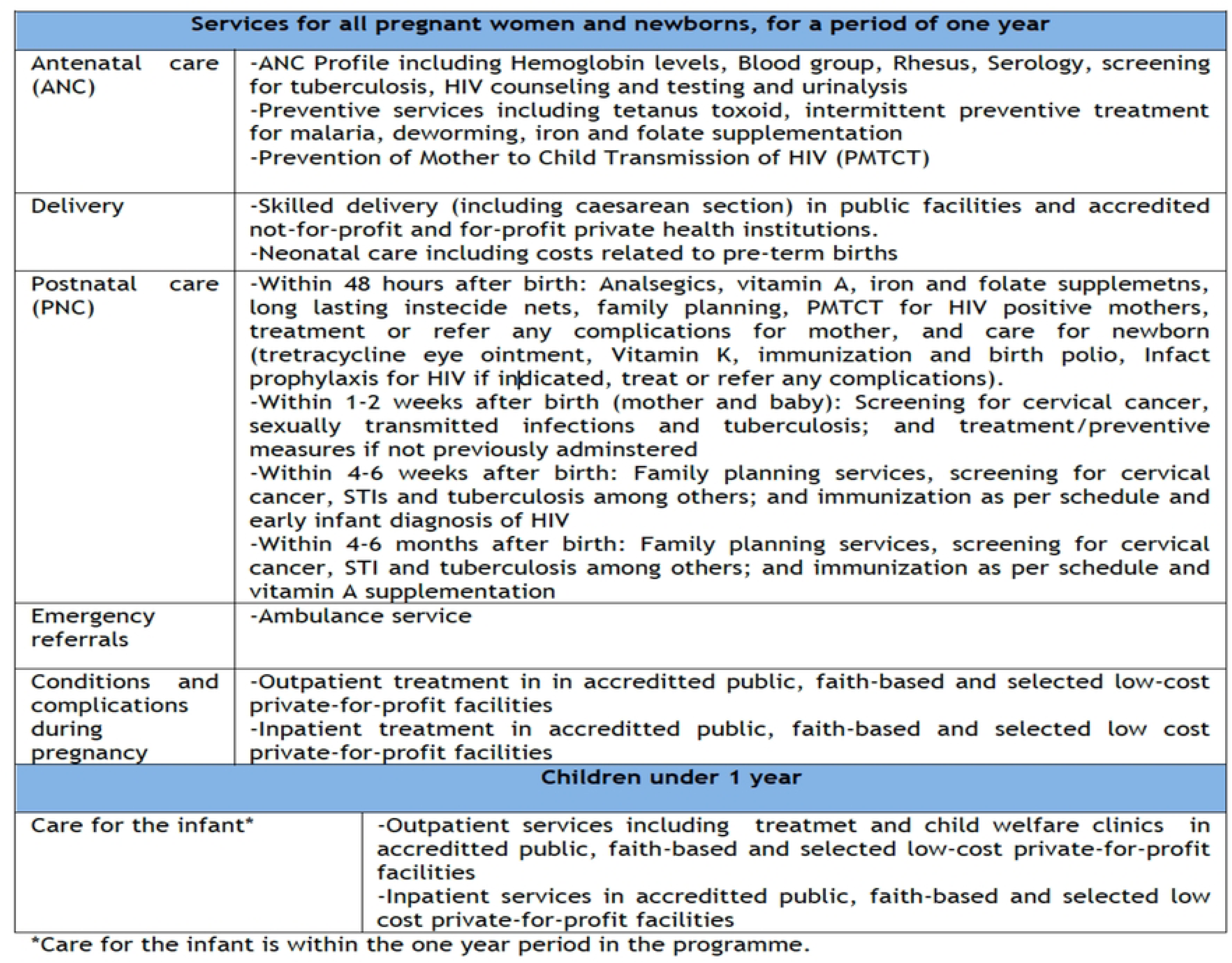
Benefit packages for Linda mama. (Source: Adopted from Implementation manual for program managers[47])

**Fig 2:**
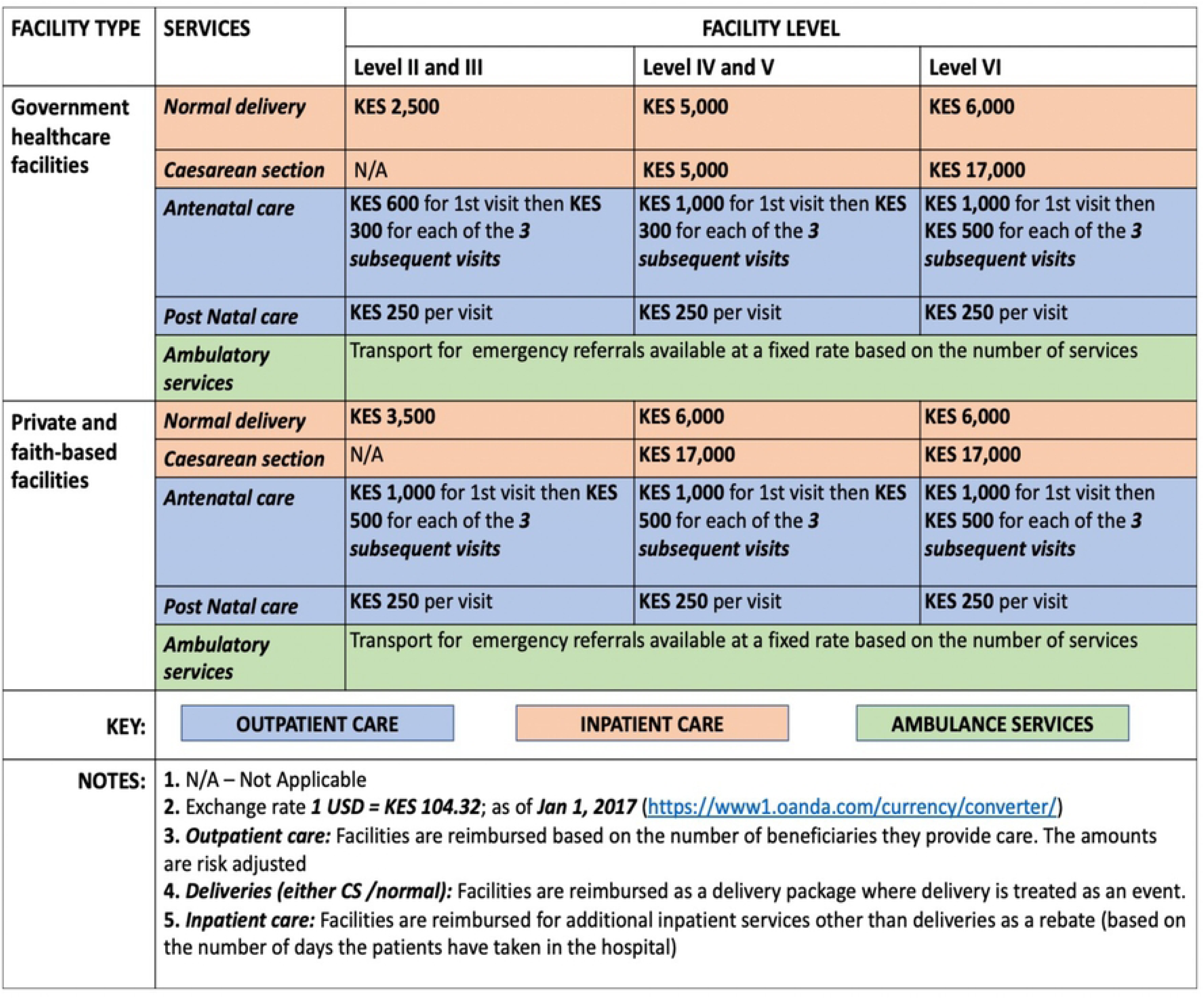
Reimbursement rates (Source: Adopted from Implementation manual for program managers[47])

### Roles of the stakeholders in the formulation and the implementation

#### A committee of stakeholders was set up to discuss the policy formulation agenda

the key informants noted that at the formulation of the current policy, the MoH set up a committee bringing together a mix of stakeholders (such as development partners, the MoH representatives, and NHIF officials) which developed a concept note (comprehensive implementation document on how Linda Mama would look like) to review the whole FM policy process and share tasks.

#### Different stakeholders played different roles in the policy formulation stage

Obtained from document reviews and IDIs, *Table 3* and *Table 5* summarises the actors who participated in the policy formulation and their roles. Characteristically, the influential participants at the formulation stage were the development partners (The World Bank, WHO, JICA, UNFPA, and USAID), who all supported the initial technical design. Of all the partners, the World Bank and JICA were most involved, as they were the co-founders of the previous FM policy. At the national level, the Presidency outlined the agenda; the appointed officials at the National Treasury allocated the budget; and the MoH, through the Principal Secretary (PS) and the director general’s (DG) office provided oversight. Interestingly, while FM policy was targeted at improving maternal and reproductive health, the members of the MoH reproductive health department felt that they were not adequately involved as noted by some respondents:

> *‘but not the team from the reproductive health stakeholders, they have not been largely involved, I mean, that is why I was saying we are all in the dark like facilities.’ – **(R023, MoH Official)***.

Equally, despite the policy aimed at improving the QoC provision of maternal care, the MoH players managing the quality and standards felt they were not adequately included as noted by some respondents:

**Table 3:** Actors roles, interest, influences and position on the formulation and implementation process of the free maternity policy.

|  | **Category of actors** | **Role in formulation** | **Role in implementation** | **Interest** | **Level of power** | **Position** |
| --- | --- | --- | --- | --- | --- | --- |
| **Elected officials** | The Presidency | + | + | High | High | Supportive |
|  | The members of parliament and senate | No | + | Low | Low | Middle support |
|  | County Governor | No | +++ | Medium | High | Middle support |
|  | Member of county assembly | No | + | High | High | Middle support |
| **Appointed officials/offices** | Office of the Auditor General | No | + | Low | Low | Middle support |
|  | Council of Governors | +++ | +++ | High | High | Supportive |
|  | The National Treasury | +++ | +++ | High | High | Supportive |
|  | Cabinet secretary, Principal secretary for health, and Director General (National) | +++ | ++ | High | High | Supportive |
|  | MoH-Department of policy, planning and health Financing (Division of Health Policy and planning and division of healthcare financing) (National) | + | + | Medium | Medium | Supportive |
|  | MoH-Department of preventative and promotive health (Division of Family Health) (National) | + | +++ | High | High | Supportive |
|  | MoH-Other departments and divisions (Standards and quality assurance and regulations, M and E) | + | + | Medium | Medium | Middle support |
|  | County Executive Committee (CEC) – Health | No | + | Medium | High | Supportive |
|  | County Chief officer of health (County) | No | +++ | High | High | Supportive |
|  | The summit ((CHMT) County directors of Health, Administration and planning and their deputies) | No | +++ | High | Medium | Supportive |
|  | The County Treasury (Includes County accountants) | No | +++ | Medium | Medium | Supportive |
|  | County NHIF focal person | No | +++ | High | Low | Supportive |
| **Purchaser of health services** | NHIF (National level) | +++ | +++ | High | High | Supportive |
|  | NHIF (County offices) | No | +++ | High | Medium | Supportive |
| **Member of interest groups** | The Church (SUPKEM, Council of churches) | + | + | Medium | High | Supportive |
|  | The Kenya Private Sector Alliance | ++ | +++ | High | High | Supportive |
|  | HCWs Unions | ++ | +++ | High | High | Supportive |
| **Donors and development partners** | The World Bank | +++ | +++ | High | High | Supportive |
|  | WHO | +++ | +++ | High | High | Supportive |
|  | JICA | +++ | +++ | High | High | Supportive |
|  | UN agencies (UNFPA) | + | ++ | High | Medium | Supportive |
|  | AMREF | + | ++ | High | High | Supportive |
|  | USAID | +++ | +++ | High | High | Supportive |
|  | Marie Stopes International | + | +++ | High | Low | Supportive |
|  | Population service International | + | +++ | High | Low | Supportive |
|  | PharmAccess | + | +++ | High | Low | Supportive |
| **Civil society** | Kenya Human Rights Commission | + | + | Low | Low | Immobilised |
|  | KELiN | + | No | Low | Low | Immobilised |
|  | Centre for Reproductive Rights | No | + | Low | Low | Middle support |
| **Beneficiaries** | Individual citizens (Men and women) | + | +++ | High | Low | Supportive |
|  | Private health facilities | +++ | +++ | High | High | Supportive |
|  | Public health facilities | + | +++ | High | Medium | Supportive |
| **Academia and researchers** | Kemri Wellcome Trust | No | ++ | High | Medium | Supportive |
|  | Population Council | No | + | Medium | Low | Middle support |
|  | Mannion Daniels and Options Consultancy | No | + | Medium | Low | Middle support |
|  | ThinkWell | No | + | High | Low | Middle support |
| **Media** | Local and international media | ++ | +++ | High | High | Supportive |
| **Other** | Beyond Zero | No | +++ | High | High | Supportive |
|  | Jacaranda Health | No | +++ | High | Low | Supportive |
|  | Philips | No | ++ | High | Low | Supportive |
|  | AfyaTu | No | + | High | Low | Supportive |
| **Key:** | +++ : Very good involvement; ++ : Good involvement; + : Partial or weak involvement; No: no evidence of involvement. |  |  |  |  |  |

Source: Author, extracted from a review of documents (**Note:** It is plausible that some actors may have been omitted because they were not apparent in the document reviews or the IDIs, KIIs, or EIs.)

**Table 4:**
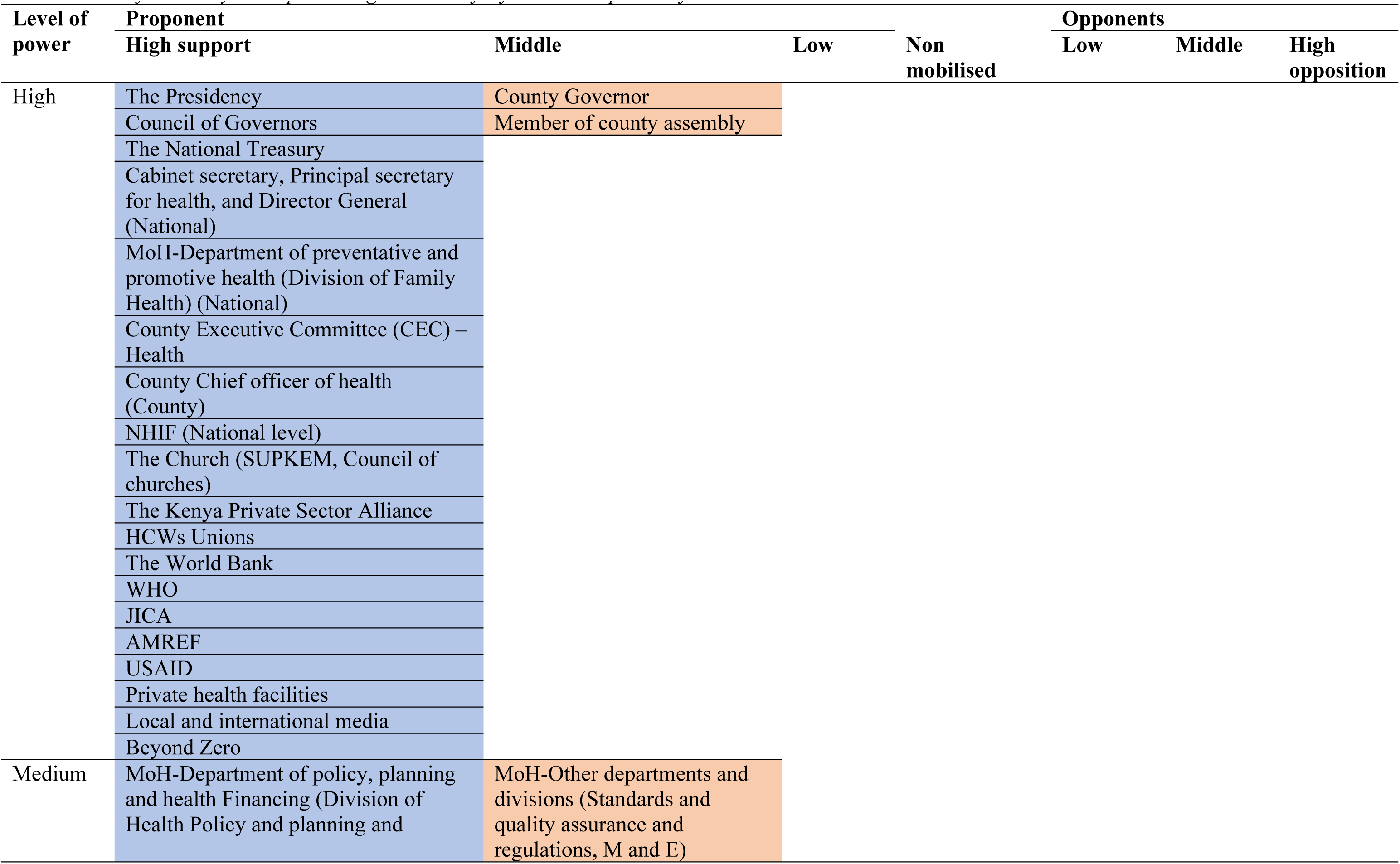
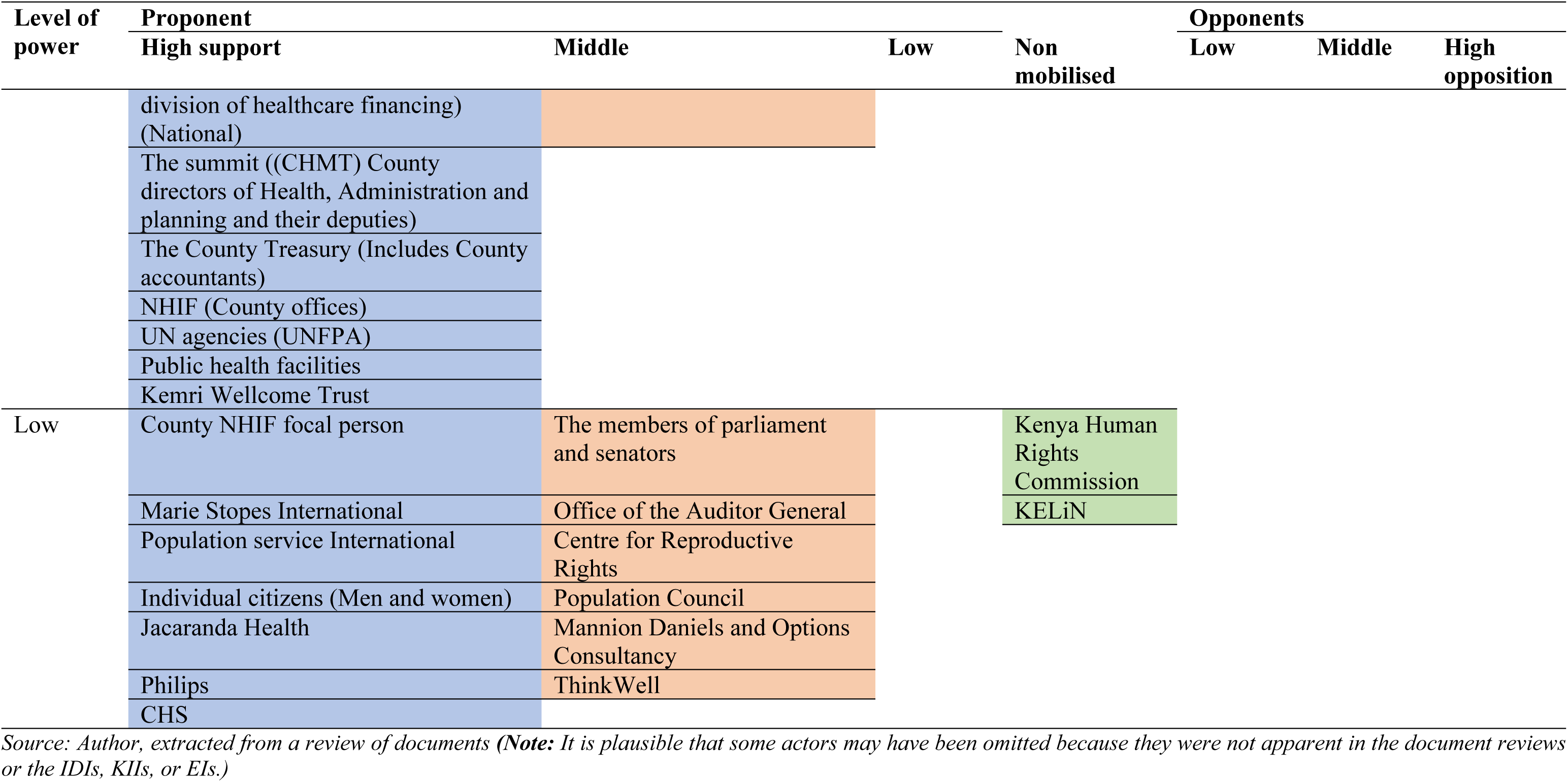
Force field analysis map showing the level of influence and power of actors.

**Table 5:**
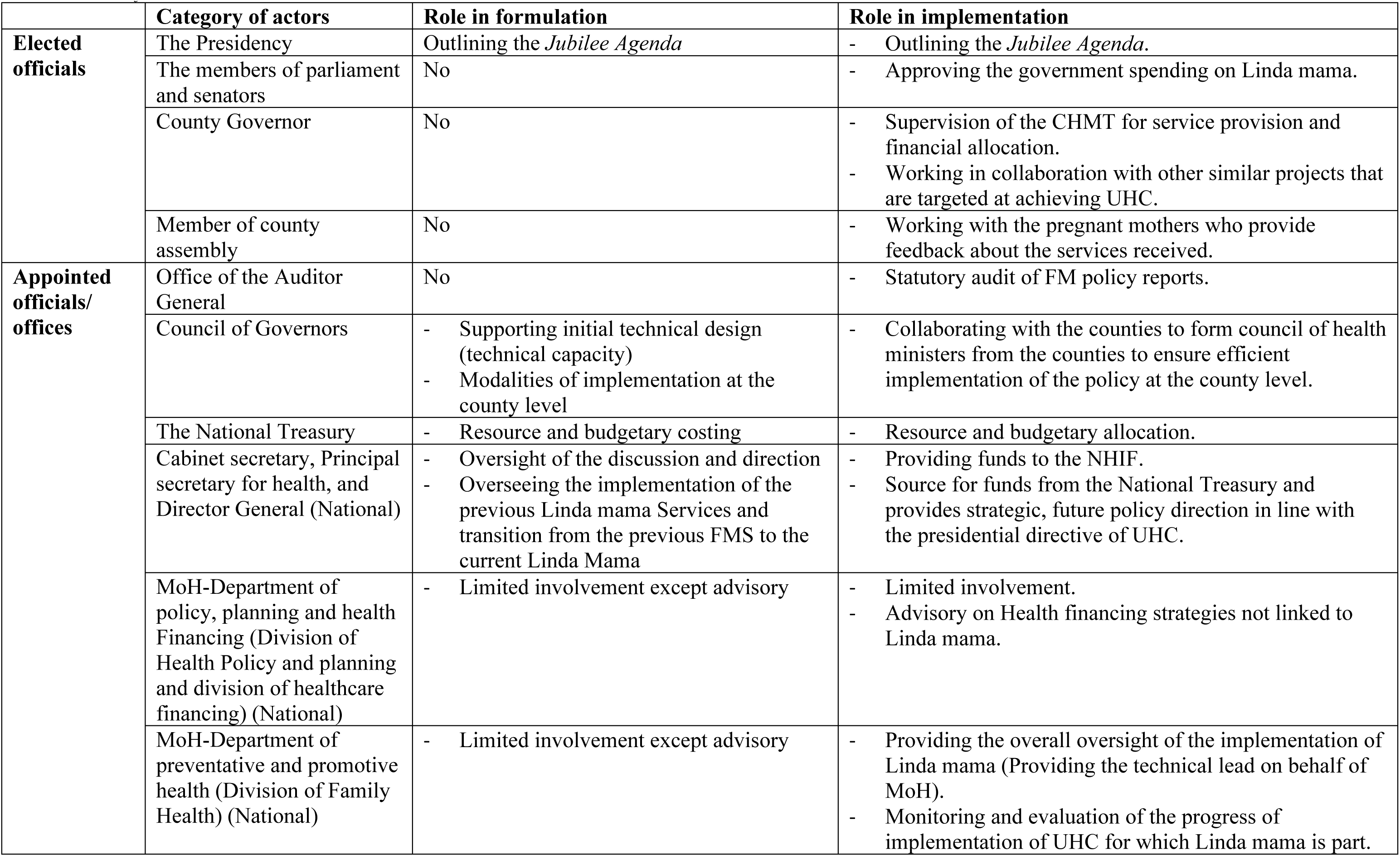
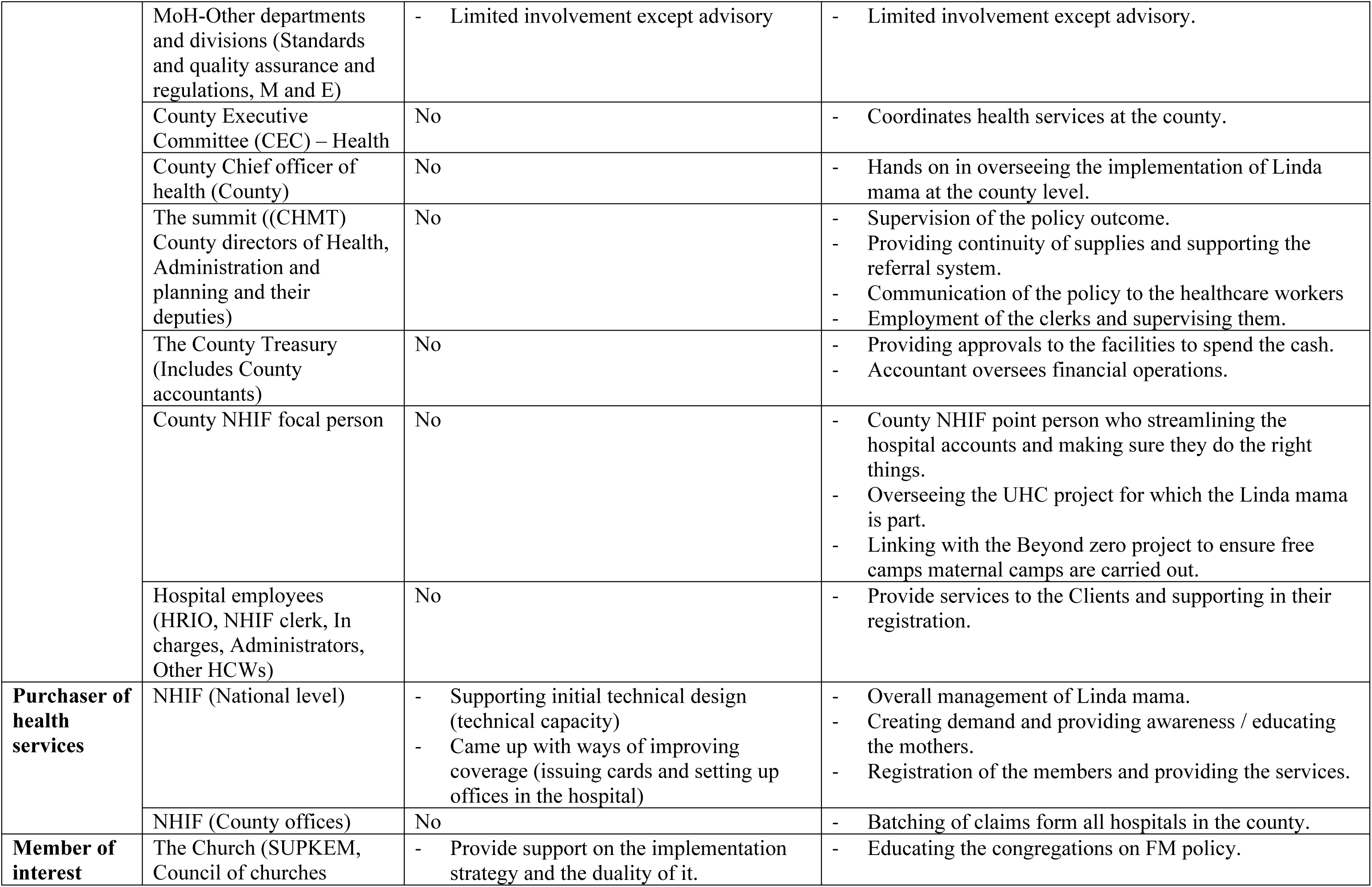
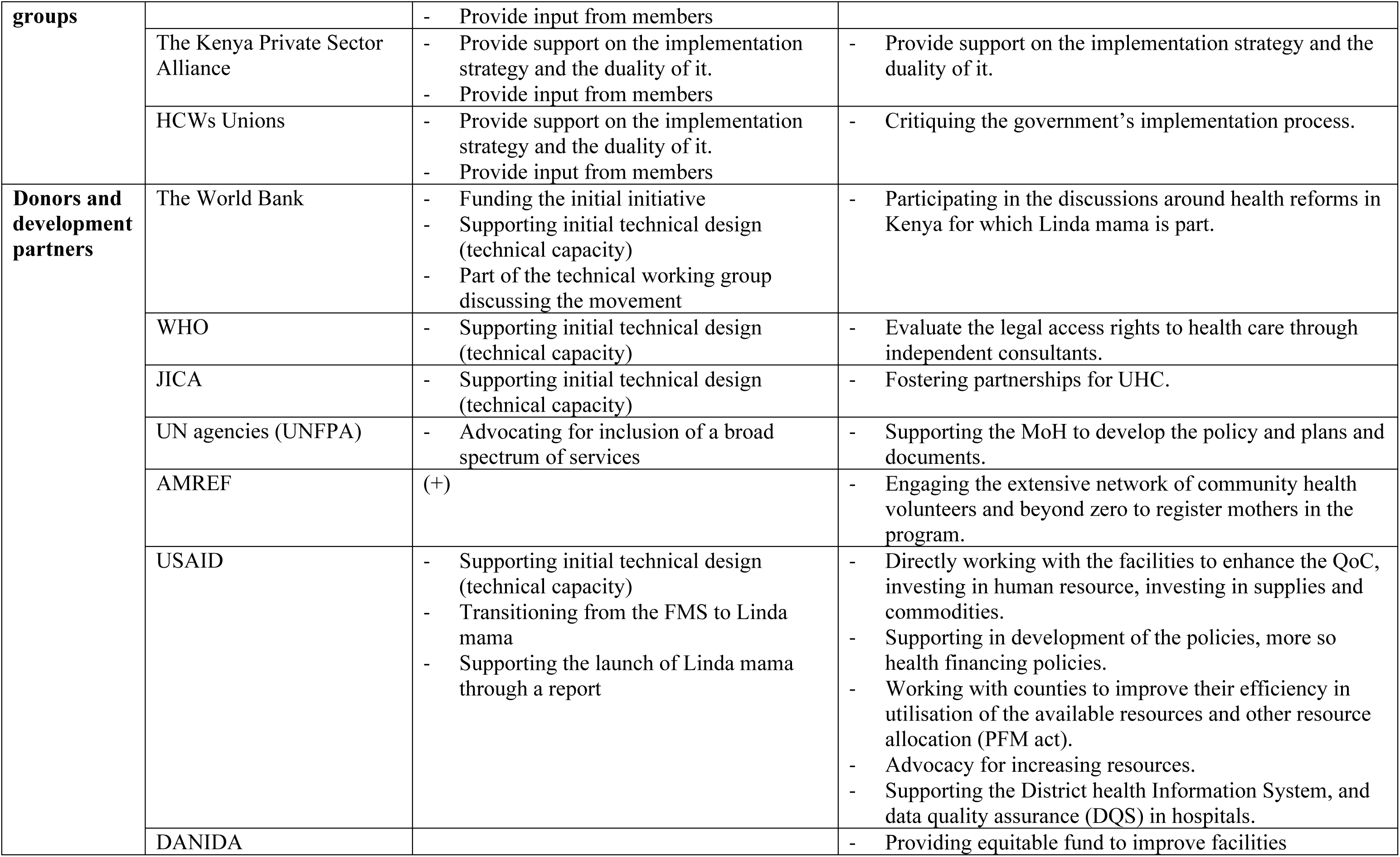
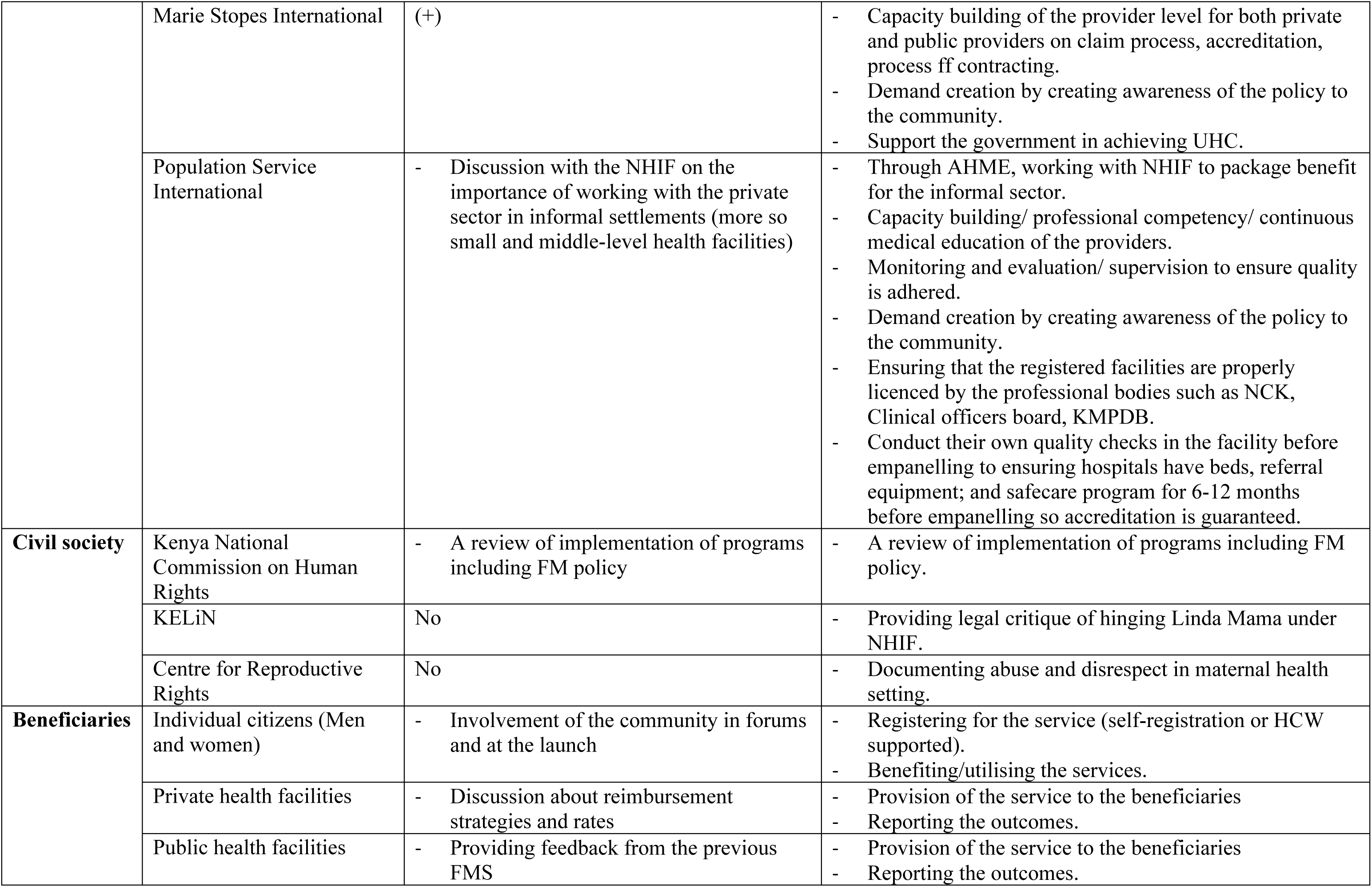
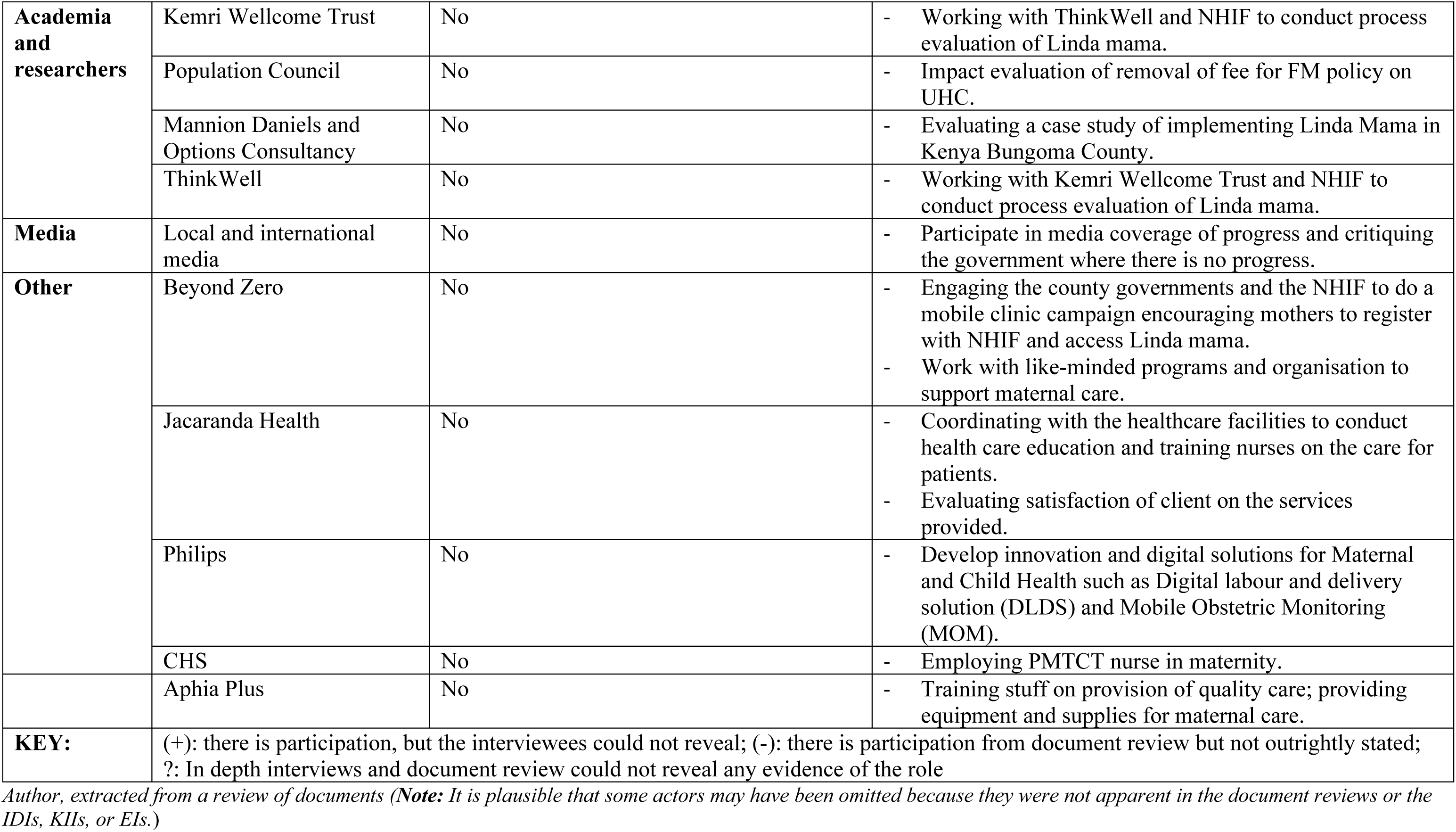
Role of the actors.

> ‘*that the players who are charged with quality just need to be roped in.’ – (**R034, MoH Official***)

The other players involved included the Council of Governors (CoG), which provided the modalities of implementation at the counties; the NHIF as the chosen purchaser of the fund, which provided implementation framework; the member of religious interest groups, private sector alliance interest group, and workers unions. Population Services International (PSI) was one of the international NGOs included in the process, which focused on persuading the committee to empanel the private sector facilities especially those they worked with to improve access in the informal settlements.

From the document review, the civil society and the beneficiaries (individual citizens and HCWs from both the private and the public sector) were involved through community forums. However, there was feeling that the mothers or their representatives, some county officials and the healthcare workers may not have been involved and did not know about the policy:

> *‘Involved? You know now, okay the change I would talk about is maybe they start involving us the people on the ground’ – **(R014, Nursing Officer)***

> *‘**I:** Were you involved in the design of the free maternity of the Linda Mama? **R:** No’ – **(R016, County level manager)***

#### There was a different organizational arrangement and role of actors in implementation

The analysis showed that implementation of the FM policy took a top-down approach in three levels: the national, county, and facility levels (Fig 3). There were more actors in the implementation than the formulation.

**Fig 3:**
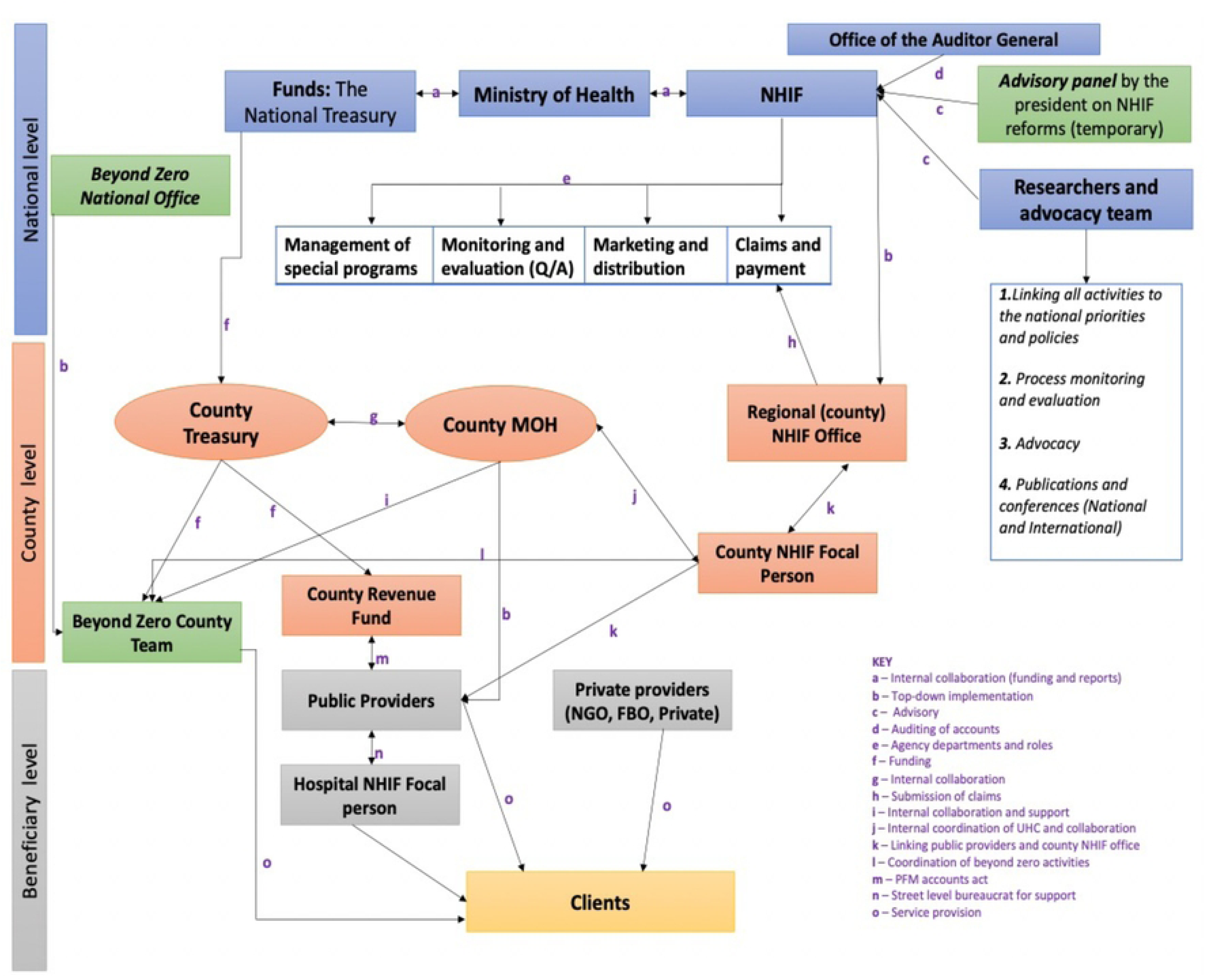
implementation arrangement of the free maternity policy as it is being implemented (source: document review and interviews)

#### In the arrangement, there was a joined-up government at the centre (national level)

The policy imperatives emerging from the national level were de facto priorities of the government as captured in the president’s *2017 Jubilee campaign Manifesto* [45]. At the national level, The Presidency, development partners, NHIF, MoH, The National Treasury, Office of the Auditor General, the CoG and the Parliament were joined up in what Exworthy and Powel [48] call ‘horizontal dimension – joined-up government at the centre’ and performed a multiplicity of implementation roles as shown in *Table 3* and *Table 5.* MoH was a powerful and influential actor at the national level. Three entities in MoH: Cabinet Secretary (CS) Health, PS health, and DG Health, were strategic policy experts, who sourced funds from the National Treasury and provided strategic, future policy direction in line with the presidential directive of UHC *(Table 4)*.

In terms of a governance structure, it was shown that it was imperative to have a proper reporting structure at the national level that would monitor the implementation of the program. However, there was a breakdown in the reporting channels that led to a gap at the national level. The effect was that the PS, who represented the MoH, would be receiving many communications concerning challenges of implementation from several sources which sometimes may have been incorrect. At MoH, the implementation was overseen by the equally powerful and influential department of preventative and promotive health (Division of Family Health), together with the NHIF, they have provided adequate social marketing to the policy through social mobilisation and communication of the providers and beneficiaries. Also, upon receiving claims and utilisation reports from the NHIF, the MoH was able to track the level of remaining funds in the pot and mobilises additional reports from the National treasury. However, the team from division of family health at MoH – despite being concerned with the reproductive health – was not involved in the formulation but passively participate in the implementation as noted by one respondent:

> *‘…but not the team from the reproductive health stakeholders, they have not been largely involved’ – **(R023, MoH Official)***.

Other departments at the MoH that were less powerful and had a medium level of interest were the division of health policy and planning, division of healthcare financing, standards and quality assurance and regulations, and monitoring and evaluation unit. The units provided strategic policy direction to the NHIF on their areas of strength and concern.

Similarly, the national treasury played a critical role but had a less influential role in the implementation process of the policy; however, they liaised with the parliament to approve the required budget for the policy. Since the introduction of the policy, they were able to provide the funds as required. Besides linking with the MOH, the National Treasury linked with the county treasury to provide other statutory funds not necessarily linked to the running of FM policy.

Equally, the NHIF was a powerful actor drawing from its mandate as an overall overseer of implementation of FM policy – as a managed fund under its department of programs and schemes – and primary purchaser of services. As a purchaser, the NHIF used its extensive network with service providers to accredit and contract providers – not previously registered on their system – for FM policy services provision. The NHIF timely reimbursed the providers for services rendered through its automated database for registration and authentication of beneficiaries. Using government ID numbers, NHIF verified the claims and redressed any complains arising from the providers on the mothers served. The NHIF had the mandate to report the claims and utilisation data for service provided, which was then yearly audited by the influential office of the auditor general. Besides, auditing the reports, the office of the auditor general was not concerned with the daily running of the implementation process.

The development partners, equally played critical roles role in the implementation such as developing financing strategies, demand generation, capacity building, and collaborations as noted by the respondents:

> *‘We do capacity building at the provider level…. it’s important that both the public and private understand the process of claim because of accreditation, process of contracting, understand issues to do with strategic purchasing in terms of …. service, how do they pay, how do they select just the whole aspect. Demand generation is one of our key aspects in terms of creation awareness.’ – **(R029, Development partner)***

> *‘We were working with NHIF to help them first of all package their informal sector product’ –* ***(R030, Development partner)***

Still, at the national level, there were two other key players: the advisory panel, researchers and advocacy team, that were less influential in the process of implementation of the policy but played an important role. The advisory panel was developed by the minister of health, albeit late in the implementation process (on 18th April 2019) in line with the Health Act of reforming and repositioning the NHIF as a strategic purchaser [49]. The team comprised the development partners, private sector, researchers, government technocrats, and advocacy coalition teams and their role was to provide the technical and financial support for NHIF, part of which is management and implementation of the NHIF. On the other hand, the researchers, mostly research institutions, and the advocacy teams, mostly the civil society, were working independently or together with the NHIF to link the activities at the county and facility levels to the national priorities, participated in the process monitoring and evaluation, provided advocacy especially of the weak and vulnerable such as adolescents, and scientific publication which were meant to improve knowledge.

#### Further in the arrangement, there was a joined-up governance at the periphery

At the county level, several players worked towards the implementation of *Linda Mama* as one respondent noted: *‘…it’s almost everyone, it’s like a teamwork’ – **(R004, Nursing Officer)**.* The two key ministries at the county that played the biggest roles in the implementation process are the treasury and health. The county treasury was concerned with receiving finance from the national treasury and providing financial support and monitoring the flow of funds at the County Revenue Fund (CRF). On the other hand, the members of the MoH at the county who oversaw the implementation of the policy form the County Health Management Team (CHMT) and were composed of several dockets such as nursing, clinical services, monitoring and evaluation, research and development, pharmacy and administration.

The dockets report to the county executive officer (CEC) of Health. Overall, the county governor [oversaw*] all the activities in the county, like especially in such free maternal its working and supervision.’ – **(R009, Nursing Officer)***.

However, as part of the CHMT, the most influential and active player in the implementation of the *Linda Mama* policy at the county was the chief officers of health as noted by the respondents:

> ‘*So now our relationship to the county is probably linked to the chief officer, through the chief officer. Because if we have issues with the implementation, then we are supposed to address them to the chief. But other people we don’t know because we don’t really see them.’ – **(R005, Facility incharge)***

The county adopted the UHC agenda of the central government by employing a county focal person for NHIF, who was important but less influential player and had a role in *‘moving forward not just with NHIF but the UHC goals of Kiambu County as a whole’ – **(R017, County level manager)**.* The regional offices of the NHIF at the county also played a significant role of receiving, batching, and quality assurance check of all the claims from the facilities in the county and sending to the national offices.

At the service provider level, there were two kinds of providers: the private and the public providers, who provided services that were responsive to needs of clients and in line with contracted terms. The public providers were part of the previous FMS that was run before while the private sector joined the service in 2017 when the new service was moved to the NHIF. They all provided the service delivery as per the benefit package and reporting of services:

*“I will say that the hierarchy and the organogram of hospital management kicks into play any time there is an issue that touches on the hospital, whether it’s Linda Mama or any other thing. We don’t have separated organs to deal with Linda Mama outside other operational issues.” – **(R010, Facility incharge)***.

Finally, the most interested but less powerful stakeholders were the beneficiaries. They were responsible for registering with the NHIF either through self-registration or HCWs assisted, utilised services and provided feedback. A summary of all roles, interests and power are in *Table 3, Table 4,* and *Table 5*.

## Discussion

This study set out to explore the process of agenda setting and policy formulation of the expanded LM policy in Kenya. Chiefly, one observation emerging from exploring the background of the policy is that it was a political initiative (guided by political campaign promises and the country’s legal and policy-guiding instruments) aimed at attaining both international (SDGs) and national goals (achieving UHC and improving access to SBA). It also sought to build on and bridge the gaps and inefficiencies of the previous FM policy implemented in 2013. This preliminary situation analysis shows that the adoption /formulation of this policy followed the opening of a ‘window of opportunity’, when a confluence of ideas and opportunities merged at the opportune time, as noted by Kingdon [50]. The confluence integrated three things: the political value of agenda-setting on such policy reforms as highlighted by Gilson et al.;[51] the need for setting the priority of policy agenda to meet national and international goals as shown by Meessen et al.;[52] and the consistency of building on FM policy that was already in the policy agenda. Further, by building on the lessons from the previous FM policy, the policy is taking the ebbs and flows fluid process instead of remaining in a fixed static form. This observation of the background of FM policy processes mirrors the practices of other countries. For instance, in Nepal, the converging interests – political and others – predestined the policy as an ideal vehicle for meeting the fortunes and objectives of the maternal incentive scheme [53].

Our findings suggest that engaging and including the private sector in the design discussion was critical as they are key stakeholders in health service provision. Sitting at the centre of the policy triangle is the issue of power, and its role in decision-making is incontrovertible. The policy formulation process was characterised by ’mixed scanning and/or muddling through.’ [15] The private sector plays a large and expanding role in healthcare service delivery, especially in sub-Saharan Africa, where the private-for-profit sector delivers 35% of outpatient care, and informal private providers deliver an additional 17% [54]. From our results, the private sector had used its power and leveraged its inclusion in implementing the policy based on its strengths of having previously developed systems such as an enhanced network of hospitals and community health volunteers and accreditation and quality monitoring standards and guidelines mirroring those of the implementing body NHIF. Researchers have shown that such network strengths (such as the developed concept of social franchise networks) fostered the public-private relationship, thereby increasing private provider accreditation into the health systems and a collegial relationship that had given small private providers more voice in the health system and improved health outcomes [55, 56].

Some interests, particularly that of setting the price for the new entrants in the policy (the private sector), were rather contentious, and the engaged representatives of the private sector had to lay the reason for the request for the higher price of reimbursements. The NGOs representing the interests of the private sector in the informal settlements devised methods to bypass the political process at the formulation to engage the sector and shape the price debate at the grassroots level rather than at the top. Through their network of influence at the grassroots level, they were able to influence the reimbursement price and the implementation. They were what Sabatier and colleagues [57–59] label as policy advocates who dominated sub-policy coalitions of actors/stakeholders. Other similar organisations have used this strategy. For instance, in the evaluation of the Africa Health Market for Equity (AHME) program, which focuses on social franchising, it was noted that the performance of the providers of the social franchise led to improved performance of *Linda Mama* policy with 79% of the social franchising facilities participating in *Linda Mama* service provision [60]. However, their contributions depend on appropriate governance prerequisites, including institutions, management capacities, and a collaborative culture to allow effective partnerships and delivery designs that target those in need and underprivileged [61].

Our findings show that the policy redesign envisaged expanding coverage and enhancing administrative efficiency, aligning with other authors’ findings [47]. The decision to make NHIF the ultimate purchaser of services was to improve the policy’s long-term sustainability and ease logistical difficulties with reimbursements, given the NHIF’s experience and reforms [62]. Further, it was to ease any legal hurdles in working with the NHIF [46]. On the other hand, while costing had been done using appropriate assumptions based on data, with the support of development partners in collaboration with Kenya’s MoH and the NHIF, it may have needed to be more collaborative, especially since some key players in MoH felt excluded. It saw the development of a costed benefits package that would be acceptable to all players and allow adequate resource allocation because it estimated crucial elements in the process. Given that the government subsidises the cost of providing health care services, especially in public health facilities (through paying for staff, medical supplies, and funding for operations and maintenance), the objective was not to reimburse the true cost of providing maternity services but to compensate for additional costs financed by women seeking care [63].

However, the private sector, NGOs and FBOs perceived their extensive investment (infrastructure, rent and staff) was high; thus, the proposed reimbursements were seen as unattractive. One study showed that government facilities have generally lower costs per service unit than Faith Based Organisations, other Nongovernmental Organisations and private-for-profit organisations (i.e., without consideration of quality, Government facilities have a higher productivity than institutions of other trustees) and that the outpatient services the spread of costs is largest for private-for-profit facilities, signifying that either the productivity of privately run outpatient services is not homogeneous, or that quality varies widely within the sub-sector [64].

The finding provides imperative lessons that with the nature in which this policy design took place, and the many conflicts and strong time pressures that were involved, a collaborative approach, in this case, was done with a long and cumbersome search for an established common ground for harnessing the difference in the design and costing without eliminating it. Due to differing interests, there is a need for deliberations and dialogue while maintaining leadership and managing conflicts at the design stage. However, this collaborative approach was essential. It saw the preparation of the policy documents related to the redesign (Cabinet Memorandum requesting cabinet approval for increased allocation to the free maternity services program, implementation guidelines, and a technical policy proposal, a communication strategy to guide the introduction of the redesigned program) [62].

The stakeholders of the *Linda Mama* policy had an opportunity to form an interaction committee or platform where the formulation design and agenda were freely discussed and debated. Researchers have shown such collaborative approaches to policy design capacitate the affected and relevant players to devise novel solutions, creating a sense of joint commitment to and responsibility for implementing the policy design [14]. Further, it facilitates a collective exploration of policy problems that allows the stakeholders to agree on novel ways of defining the problem that both emphasise its urgency and make it solvable. Noteworthy is Dye’s [65] assertion that public policies often reflect the interests, preferences, and the values of the governing elites were evident in the committee. While the Presidency was not an active participant in the committee, the technocrats had to align the design to fit into his political promise and agenda of achieving the UHC. However, the participation of actors in the policy formulation was not as inclusive. Grindle and Thomas’s [66] observation that despite the characteristics of policy actors such as personal attributes, loyalties, institutional and political commitments, and training, the actors are never completely autonomous. The stakeholders had to work within a meshed context and had to tackle the problems and issues (policy formulation) that they faced and provide a well-thought-through solution that was economically, politically, and administratively feasible.

The appointed government officials (often the technocrats in government), the development partners, and the representatives of the beneficiaries had a substantial influence on the formulation of the details of the policy. Whereas there were representatives of the beneficiaries, the results showed the beneficiaries interviewed did not know about any form of public participation. While it would be impractical to include all the participants in the formulation, the level of public participation in reforms highlights Grindle and Thomas’s [66] assertion that the participation is a determined categorisation of the reform as a bureaucratic compliance reform, that requires limited or ‘invisible’ public participation or as requiring political stability and support which need ‘visible’ participation or a comprehensive public engagement. The finding shows that the beneficiaries’ representatives were significant because they were classified in the FM reform as the latter. Only representatives voiced the participants’ interests. In fact, in the formulation process of FM policy in Ghana, authors categorised the participants as policy agenda directors, agenda approvers, government and non-government agenda advisors, and agenda advocates [16]. The agenda advocates, including the beneficiaries, are active throughout the formulation process. Generally, there was good coordination of the policy processes at the formulation level; but, if the beneficiaries and the implementers are not well involved at the policy design stage, it would bring into focus the question of how committed they were to implement things that they had little say.

Significantly, more actors at the implementation level – both at the national and the county level – supported achieving the goals in audit, research, financing and strategic operations. These roles by different actors affirm the findings in other research that actors look for the mobilisation of specific capacities to address particular policy problems, in this case, supporting policy implementation [67]. Implementers of the *Linda Mama* policy understood their roles as was formulated; however, the communication at the national level was rather complex and messier since there was a gap in clearly defined roles in the communication network at the MoH. Every department wanted an ‘ear’ of the influential PS health in providing strategic decisions. Besides, the dependency of the policy imperatives at the top level was significantly higher as units had to rely on one another more often than necessary, slowing down the implementation cascade. These findings affirm that the *Linda Mama* policy has achieved three of the ten proposals Hogwood and Gunn [68] postulated for successful implementation. However, there is an interaction and better coordination of roles at the periphery (county and facility level) in that the county has gone further to employ someone to ease communication and coordination with different national and county departments. While there are gaps in the implementation perspectives, our work has captured the importance of implementation readiness of policy, which is done by ensuring that organisations and players responsible for implementation have accepted and support the legitimacy of the policy and that there is sustained political support which comes with enhanced policy clarity of the objectives and that players have all these set within their local contexts [69–71].

Overall, our findings are consistent with the exploration of the FM policy formulation literature elsewhere,[72] decision agendas of the *Linda Mama* policy were driven by a complex interplay of factors related to the context, processes and content, actors and their interests, power and roles, in an elaborate manner rather than linear processes; thus, reiterating the enduring relevance and validity of Walt and Gilson’s policy triangle [27]. This finding captures well Walt et al.’s [73] statement that *‘there are also many other conceptual challenges…capturing and measuring the level of resources, values, beliefs and power of diverse actors is difficult.’*

## Conclusion

The policy formulation process and the agenda-setting of the Kenyan free maternity policy presented drivers through policy content and ideas, context, actors and their powers, and issue framing within both bureaucratic and public venues to better understand the policy process. Our study highlights the basis for changing the previous policy to the current one driven by multiple agendas, including political determinants, the need to achieve global and national goals, and learnings from the previous policy. The interconnectedness of these drivers shaped the content and policy formulation processes. Additionally, it shows the influence of an actor’s power on the policy process depends on the actor’s interests, ideas and position to mobilise the agenda decisions to move in specific directions. Understanding policy processes is relevant across other LMICs, and we hope that this framework contributes to policy analysis and learning in Kenya and beyond.

## Data Availability

All data generated or analysed during this study are included in this published article as quotations. The original qualitative data are not made available since some responses contain potentially identifiable information at multiple levels, including for individuals, health facilities, and organisations. The Institutional Review Board conditions stipulated that participants would not be at risk of identification through their responses participant consent forms did not include obtaining explicit consent regarding the possible use of anonymised data in the public domain via a data repository. For non-author contact information for enquiries about data access, please

## Acknowledgements

The authors acknowledge all those who participated in the study and donated their time and skills.

## Notes

### Competing Interest Statement

The authors have declared no competing interest.

### Funding Statement

This study was funded by the Commonwealth Scholarship Commission [KECS-2017-266], which supported BO's PhD study. The funding agency did not play any role in the analysis, interpretation of results, and manuscript writing.

### Author Declarations

Ethical approval for this study was obtained from the University of Kent, the SSPSSR Students Ethics Committee and the AMREF Scientific and Ethics Review Unit in Kenya (Ref: AMREF ESRC P537/2018). Further, we received permission to conduct the study from all the healthcare facilities where the study was conducted and additional clearance to conduct research from the County Government of Kiambu, Department of Health Services (Ref. No: KIAMBU/HRDU/AUTHO/2018/10/31/Oyugi B). We also got written informed consent from the respondents before conducting the interviews, after informing them about the purpose of the study and their right to withdraw consent at any point. They were also assured of their confidentiality, and that their data would be reported in an aggregated format, and anonymised to protect their identities, throughout the course of this study.

